# Large-scale web scraping for problem gambling research: a case study of COVID-19 lockdown effects in Germany

**DOI:** 10.1101/2022.07.27.22277642

**Authors:** Elke Smith, Simon Michalski, Kilian H. K. Knauth, Kai Kaspar, Nils Reiter, Jan Peters

## Abstract

The COVID-19 pandemic and the measures to prevent its spread have had a negative impact on substance use behaviour and posed a special threat for individuals at risk. Problem gambling is a major public health concern, and it is likely that the lockdown and social distancing measures have altered gambling behaviour, for instance shifting from land-based to online gambling. In this study, we used large-scale web scraping to analyse posting behaviour on a major German online gambling forum, gathering a database of more than 200k posts. We examined the relative usage of different subforums, i.e. terrestrial, online gambling and problem gambling sections, posting frequency, and changes in posting behaviour related to the casino closures that were part of the nationwide restrictions in Germany in 2020. There was a marked increase in the number of newly registered users during the first lockdown compared to the weeks prior to the lockdown, which may reflect a shift from terrestrial to online gambling. Further, there was an increase in the number of posts in the online gambling subforum with a concurrent decrease in the number of posts in the terrestrial gambling subforum. An analysis of user types revealed that a substantial number of users who posted in both the online and terrestrial forum contributed at least once to the problem gambling subforum. This subforum contained the longest posts, which were on average twice as long as the average post. Modelling the relationship between reply frequency and latency between initial posts and replies showed that the number of short-latency replies (i.e. replies posted within seven hours after the initial post) was substantially higher during the first lockdown compared to the preceding weeks. The increase during the first lockdown may reflect the general marked increase in screen time and/or usage of online platforms and media after the onset of the global COVID-19 pandemic. The analyses may help to identify lockdown-related effects on gambling behaviour. These potentially detrimental effects on mental health, including addiction and problem gambling, may require monitoring and special public health measures.

## 1 Introduction

The rise of social media use in conjunction with advances in machine learning, text mining and modelling have created novel opportunities for the use of social media “big data” to address research questions in psychology (Conway & O’Connor, 2016; Landers, Brusso, Cavanaugh, & Collmus, 2016). One area that has attracted particular attention concerns the association between mental health problems and social media behaviour, e.g., with respect to the type of content posted on social media sites (Chancellor & De Choudhury, 2020; Merchant et al., 2019). In this line of work, for example, substance use was linked to posting behaviour on Twitter^©^ (Cavazos-Rehg et al., 2015; Glowacki, Wilcox, & Glowacki, 2021; Lamy et al., 2016). Also, some studies analysed gambling-related content on Twitter^©^ created by online gambling operators (Killick & Griffiths, 2020) and users (Fino, Hanna-Khalil, & Griffiths, 2021). While general social media sites such as Twitter^©^ continue to be widely used, more extensive discussions of specific and potentially problematic activities (such as gambling) often occur in focused online communities. In the context of the COVID-19 pandemic, social media data have been leveraged to examine pandemic-related mental health problems and challenges, for instance with respect to addiction (Glowacki et al., 2021) and gambling behaviour more specifically (Fino et al., 2021).

### The COVID-19 pandemic and gambling behaviour

The worldwide spread of the coronavirus SARS-CoV-2 starting in December 2019 and the resulting COVID-19 pandemic constitutes a substantial psychological stressor: due to repeated lockdowns, social interactions and everyday behaviours are affected and regulated in unprecedented ways (Wu et al., 2022). This is paired with high uncertainty regarding how the pandemic will evolve and how it will affect individuals. School and day care closures and job insecurities put additional strain on families (Martinsone & Tzivian, 2021). Accordingly, early meta-analyses suggest high levels of COVID-19-related psychological stress (Cooke, Eirich, Racine, & Madigan, 2020). The COVID-19 pandemic and the associated containment measures put in place have had a negative impact on substance use behaviour (Boschuetz, Cheng, Mei, & Loy, 2020; Killgore, Cloonan, Taylor, Lucas, & Dailey, 2021; Vanderbruggen et al., 2020), very likely a consequence of heightened stress, social isolation and feelings of loneliness. It is reasonable to assume that this might also apply to gambling behaviour. In substance use disorders, stress is known to trigger craving and relapse (Hellberg, Russell, & Robinson, 2019; Mantsch, Baker, Funk, Lê, & Shaham, 2016; Sinha, Sinha, Li, Sinha, & Li, 2007). Along similar lines, the severity of problem gambling is associated with stress (Elman, Tschibelu, & Borsook, 2010; Loo, Oei, & Raylu, 2011).

The public on Twitter views the COVID-19 pandemic as a special threat to certain individuals, being more exposed to online gambling content (Fino et al., 2021). In the context of leisure activities, there was a marked change in the use of communication and social media during the lockdown periods (Meier, Noel, & Kaspar, 2021). COVID-19 and the lockdown and social distancing measures put in place may therefore also alter gambling behaviours (e.g., behaviour might shift from terrestrial to online gambling). A study from a Canadian sample found that some individuals switched from terrestrial to online gambling during the COVID-19 pandemic and that especially high-risk gamblers exhibited an increased likelihood of online gambling behaviour (Price, 2020). An increase in online gambling during the COVID-19 quarantine period was reported in a dataset from an online study with mainly UK and US participants (Sallie, Ritou, Bowden-Jones, & Voon, 2021). In contrast, reduced online gambling behaviour (online sports betting) was reported after COVID-19 measures were put in place for a dataset from Sweden, Germany, Finland, and Norway (Auer, Malischnig, & Griffiths, 2020). A study from an American sample observed reduced online gambling, but increased substance use (Xuereb, Kim, Clark, & Wohl, 2021). In that study, only a minority of individuals who gambled substituted terrestrial with online gambling. However, these appeared to be vulnerable individuals with symptoms of problematic gambling behaviour. In a sample of Swiss terrestrial gamblers, no changes in online gambling participation during lockdown in Switzerland were found, but those who gambled online during lockdown played more often and longer than before (Lischer, Steffen, Schwarz, & Mathys, 2021).

Changes in gambling behaviour caused by COVID-19 and the measures put in place have not been extensively studied to date and appear to heavily depend on the population or type of gambling activity studied. The effects of the COVID-19 pandemic on gambling behaviour likely depend on complex local and individual factors, including shutdowns of gambling venues and limited availability of sports betting due to cancellation of events (Auer et al., 2020; Gainsbury, Swanton, Burgess, & Blaszczynski, 2021). In Germany, terrestrial gambling facilities have remained closed over longer periods of time due to lockdowns, which may have led to a shift from terrestrial to online gambling in certain individuals (see Supplement, Table S1). Importantly, the vast majority of the studies cited above rely on self-reports to quantify changes in gambling behaviour associated with COVID-19 measures. However, self-reports are subject to various biases, including social desirability (Krumpal, 2013) and recall bias (Bradburn, Rips, & Shevell, 1987), whereby computer-based surveys appear to yield significantly more reporting of socially undesirable behaviours compared to equivalent paper-based surveys (Gnambs & Kaspar, 2015). Therefore, the field might benefit from more objective measures of changes in gambling behaviour associated with different societal stressors or policy changes.

### Gambling-specific online communities

Online gambling communities typically constitute discussion boards or internet forums where users share or discuss gambling experiences, strategies or gambling problems (Griffiths, 2010; Sirola et al., 2021). Survey studies revealed that high levels of engagement in online gambling communities occur more frequently in players suffering from higher levels of problematic gambling behaviour (Sirola, Kaakinen, & Oksanen, 2018; Sirola, Kaakinen, Savolainen, & Oksanen, 2019). For example, amongst participants in a survey amongst Finnish gamblers (Sirola et al., 2018), 54.33% of online gambling forum users reported South Oaks Gambling Screen (SOGS) (Lesieur & Blume, 1987) scores *>* 2 (“some problems with gambling”), whereas this was the case for only 15.58% of respondents who reported to have never visited such a community site (Sirola et al., 2018). Studying such online communities might therefore yield insights into problematic gambling behaviour (Griffiths, 2010).

Engagement in online (gambling) communities can be investigated using a variety of approaches. On the one hand, individual posts can be subjected to detailed manual content analysis (Bourgonjon, Vandermeersche, De Wever, Soetaert, & Valcke, 2016). This can yield important insights into the specific issues and themes that are discussed in a given community. However, due to technological advances, very large data sets can now be automatically retrieved (scraped) from the web (Landers et al., 2016). For such data, manual content analyses are clearly not feasible. At least two alternative approaches are therefore possible. First, one could instead focus on posting behaviour, rather than post content. The former is easily quantifiable in large data sets, e.g. in terms of the number of posts or active users in a given time window or a specific subforum of a community, and might yield novel insights. Second, one could use data-driven text mining approaches such as topic modelling (Blei, Ng, & Jordan, 2003) to investigate the prevalence of different topics in online communication (Carron-Arthur, Reynolds, Bennett, Bennett, & Griffiths, 2016; Fino et al., 2021). In the present study, we used large-scale web scraping to carry out a detailed analysis of posting behaviour for a large German online gambling forum. By focusing on objective online behaviour (e.g., posting behaviour with respect to a dedicated gambling forum), we circumvented known shortcomings of self-reports, such as the social-desirability bias (Krumpal, 2013) and recall bias (Bradburn et al., 1987). Specifically, we examined the relative use of different subforums, overall posting frequency, and event-related changes in posting behaviour, with a total data base of *>* 200k posts. We first describe the data set in detail. In a second step, we report analyses of posting behaviour related to the casino closures that were part of the nationwide restrictions in Germany to prevent the spread of COVID-19 in 2020. During the first lockdown period in Germany starting in March 2020, all “non-essential” facilities and businesses were closed, including schools and universities, restaurants, bars, clubs, and casinos. Citizens were asked to reduce social contacts to an absolute minimum and spending time in public was only permitted for small groups. A period with reduced incidence rates and re-openings in summer 2020 was followed by rising incidence rates of COVID-19 in fall 2020, and a second lockdown was instated in November, during which casinos and other “non-essential” facilities were closed again. We hypothesised that changes in gambling behaviour would be reflected in the statistical properties of the posts, such as frequency and distribution across the subforums.

## 2 Methods

### 2.1 Description of the discussion board

The discussion board is part of a German-speaking online casino and gambling website. We use the following terms to describe the discussion board: A *post* is a single text from an individual user. A *user* is a person with an account (profile page) who may write posts. An *initial post* is a post that is not a response to another post. A *reply* is a post that is created in response to another post. A *thread* constitutes an initial post plus all of its replies. Threads are grouped in eleven superordinate board topics, which centre around online casinos, slot machines, games such as roulette, blackjack and poker, gambling and gambling addiction. In order to compare posts related to online and terrestrial gambling, only subforums and, in some cases, individual subcategories of subforums that clearly state in their description that only online or terrestrial gambling topics may be posted there, are considered for the analyses. The *online subforum* therefore contains exclusively online gambling sections and the *terrestrial subforum* exclusively terrestrial gambling sections. Sections of the forum for which it is unclear whether the content is associated with online or terrestrial gambling are therefore not included in the online and terrestrial subforum, respectively. The *problem gambling subforum* refers to the problem gambling board topic and does not overlap with the online or terrestrial subforums. This subforum was included in the analysis due to the potential clinical relevance of its use.

A single post consists of a headline and content. Each post has a publication date, an author (user name), a designated position in the thread, a count of likes, and an indication whether the post constitutes a reply to another post. Posts may also be empty (post deleted if the content contained insults or topic closed by the board administrators).

A *regular* user has a unique profile page. A user may also be flagged as *blocked, anonymous* or *deleted*, in which case no unique user name or profile page is displayed or accessible. A single user has a unique author name, a status (e.g., *starter* and *verified* user), a count of posts, and a date of joining the discussion board. Newly registered users have the status *starter* for a certain period of time, depending on their activity. To become a verified user, a photo of the face and a sheet of paper with the user name and date must be uploaded.

We classified the users as belonging to one of the following user types: *Online only* (at least one post in the online gambling subforum and zero posts in other sections of the forum), *terrestrial only* (at least one post in the terrestrial gambling subforum and zero posts in other sections of the forum), *mixed* (at least one post in the online gambling subforum, at least one post in the terrestrial gambling subforum and zero posts in the problem gambling subforum), *online only + PG* (same as online only, but additionally at least one post in the problem gambling subforum), *terrestrial only + PG* (same as terrestrial only but additionally at least one post in the problem gambling subforum), *mixed + PG* (same as mixed but additionally at least one post in the problem gambling subforum) and *PG only* (at least one post in the problem gambling subforum and zero posts in other sections of the forum). Users that do not fit in any of the above categories are classified as *other*.

### 2.2 Data collection

The website was scraped in April 2021 using *Python* (version 3.6.9) and the *Requests* (version 2.18.4) and *Beautiful Soup* (version 4.6.0) libraries for *Python*. The obtained XML data were parsed into a relational *SQLite* database using *Python* and the *sqlite3* module for *Python*.

### 2.3 Legal aspects

Web scraping for scientific purposes is generally legal, as long as no access restrictions are circumvented (Klawonn, 2019). While users hold copyright to their posts, the German Copyright Act *UrhG* allows text and data mining. The data scraped from the website were generally accessible and technical measures designed to prevent web scraping were not disregarded. The scraped public user profiles do not contain personal information that would allow an individual person to be identified. The research conducted on the basis of the data serves non-commercial purposes only and the published results do not allow identification of natural persons.

### 2.4 Definitions of the lockdown periods

Closures of gambling halls in the federated states of Germany commenced between March 14 and 19 in 2020. Venues re-opened between May 4 and June 15th in 2020. The first lockdown was therefore defined as the period from Monday, March 16 to Sunday, May 3 of 2020, to enable an analysis of whole weeks. The first pre-lockdown period corresponds to the seven weeks before the first lockdown period. The second lockdown was then defined as the period from November 2 of 2020 to December 20 of 2020, to obtain a period of equal length as the first lockdown (the gambling halls were simultaneously closed down on November 2 of 2020 in all federated states of Germany and reopened in spring 2021). The second pre-lockdown period corresponds to the seven weeks before the second lockdown period. The lockdown phases in Germany in 2020 are depicted in Figure 1, for an overview on the casino closure periods in the federated states of Germany, see Supplement, Table S1.

**Figure 1:**
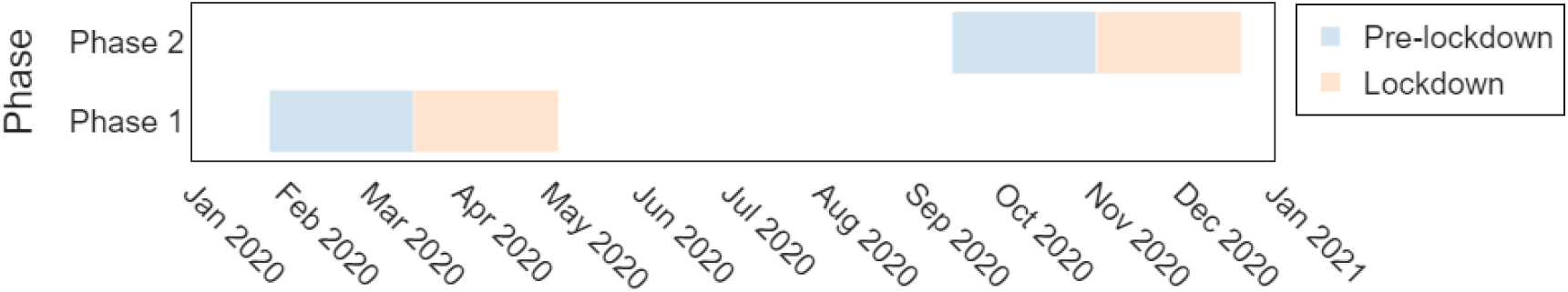
COVID-19 lockdown phases in Germany in 2020. The pre-lockdown periods are defined as the seven weeks before the respective lockdown phases.

### 2.5 Modelling of reply latencies

User engagement may not only be reflected in the overall number of posts and replies, but also in the timing of user behaviour. To address this issue, for each of the four pre-defined periods (prelockdown 1, lockdown 1, pre-lockdown 2, and lockdown 2), we modelled the temporal evolution of reply frequencies following an initial post. We only included replies within the first 7 days following the initial post, and only considered initial posts up to seven days before the end of the respective period to ensure that all considered reply posts fall within the respective period. The reply latencies were binned into bins of 8 hours (resulting in 21 bins in total, ranging from 0 to 167 hours for a complete week). We modelled the reply latencies as exponential decay with

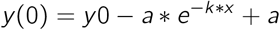

were *y0* is the intercept, *a* the asymptote and *k* the decay rate. The parameter distributions were estimated via *Hamiltonian Monte Carlo* sampling (4 chains, 4000 samples, warmup = 2000, thinning = 2) using *Stan* (2.21.0), and the *Python* interface to *Stan PyStan* (2.19.1.1). Chain convergence was determined such that 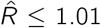 (Gelman & Rubin, 1992). We report directional Bayes factors (*dBFs*) (Kass & Raftery, 1995; Marsman & Wagenmakers, 2017) to quantify the degree of evidence for reductions and increases, respectively, in parameter values for *y0, a* and *k* for the pre-lockdown and lockdown phases via kernel density estimation in Python. The *dBFs* are defined as the ratio of the integral of the posterior distribution from - ∞ to 0 versus the integral from 0 to ∞. For instance, a *dBF* of 10 indicates that a positive directional effect is ten times more likely than a negative directional effect.

## 3 Results

We first describe the dataset, before focusing on the specific analyses of COVID-19 lockdown-related effects.

### 3.1 Characteristics of the discussion board

The discussion board is divided into eleven superordinate board topics. In total, there were 205,385 single posts, pertaining to 7,902 threads and 4,428 registered user profiles (excluding anonymous and blocked users whose number cannot be determined). Anonymous and blocked users wrote 46,100 of the 205,385 posts.

#### 3.1.1 Users

On average, 36.00 days (*SD* = 125.24) passed between registration on the platform and the first post. 1552 users were flagged as *starter* (35.05% of all users), and 181 as *verified* users (4.09% of all users). There were on average 35.94 posts per user (*SD* = 168.81). Most users (3272 users or 74% of all users) published between 0 and 10 posts (see Figure 2A). Many users posted only once (*N* = 1214; 27.42% of all users), or wrote several posts within a day (*N* = 751; 16.96% of all users), and then no more. However, there are also many users who are active in the forum over a longer period of time, for which the time between the first and the last post is one year (*N* = 867; 19.58% of all users) or more (*N* = 802; 18.11% of all users) (see Figure 2B). The 100 most active users (2.26% of all users) contributed to 42.92% of the posts. There was a marked increase in newly registered users during the first lockdown phase in 2020 (see Figure 2C). There were 175 new registrations during the first pre-lockdown phase in 2020, in comparison to 281 new registrations during the first lockdown phase, which represents an increase of 60.57%. During the second pre-lockdown period there were 199 new registrations, compared to 215 new users during the second lockdown period (increase of 8.04%). A large proportion of users were classified as *online only*, i.e. had at least posted once the online gambling subforum, but not in other subforums (see Figure 2D). About half of the users did not fit into any of the predefined categories (classified as *other*, see section 2.1). Among the less frequent user types, the most frequent category was *mixed + PG*. Only very few users posted exclusively in the problem gambling subforum or terrestrial gambling subforum.

**Figure 2:**
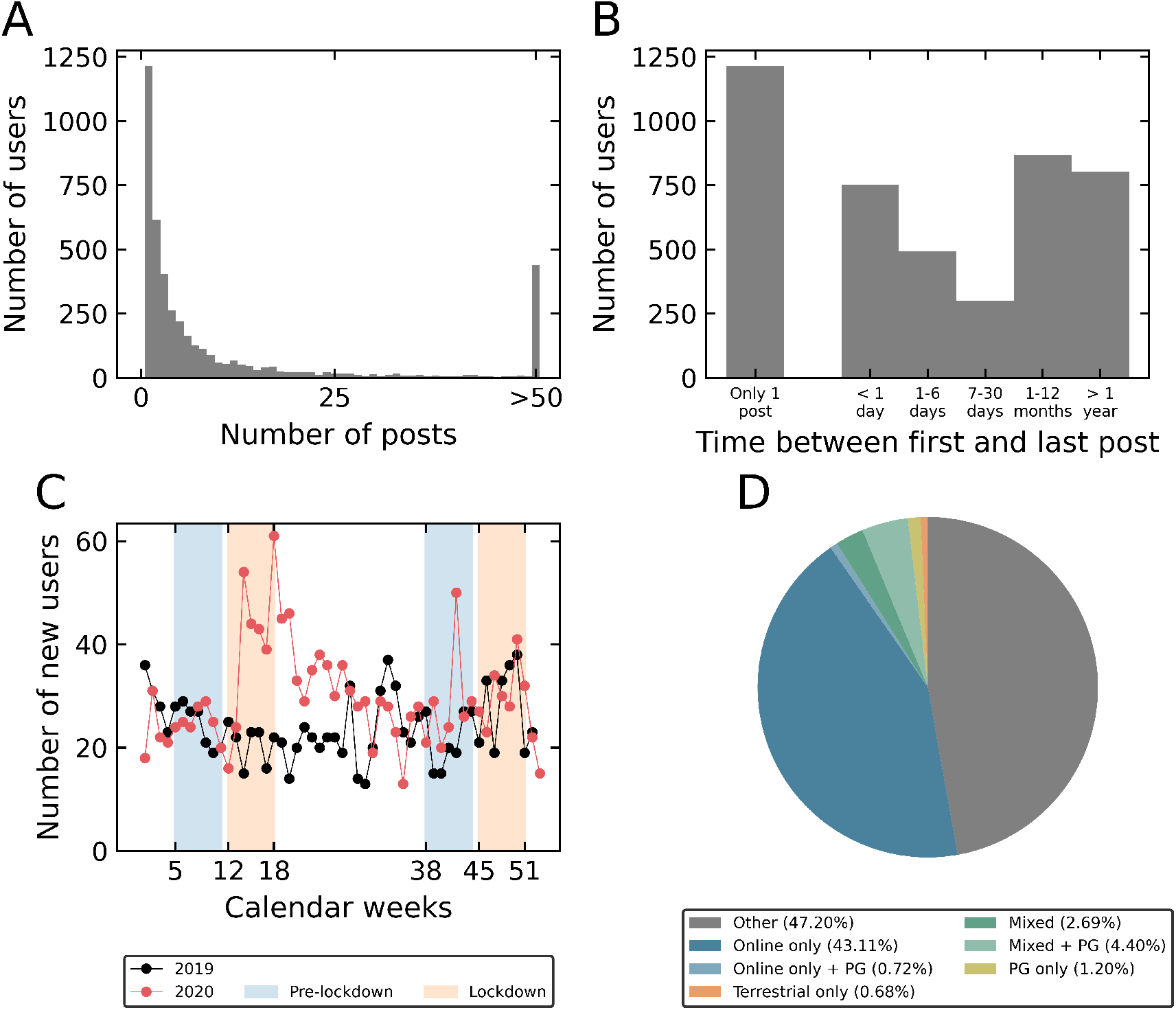
A: Distribution of posts and users; B: Time between first and last post; C: Number of newly registered users per year, calendar week and phase (pre-lockdown and lockdown), D: Proportion of user types. Zero users were classified as *terrestrial only* or *terrestrial + PG. PG* = problem gambling.

#### 3.1.2 Posts

The online gambling subforum contained the most posts (*N* = 135,785; 66.11% of all posts), followed by the problem gambling subforum (*N* = 4150; 2.02% of all posts), and finally the terrestrial gambling subforum (*N* = 3365; 1.64% of all posts). The percentage of initial compared to reply posts was similar for the subforums (42.78% to 57.22% for the entire forum, 45.03% to 54.97% for the online gambling subforum, 41.60% to 58.40% for the terrestrial gambling subforum, and 41.40% to 58.60% for the problem gambling subforum). During the first lockdown compared to prelockdown period, the number of posts increased by 41.48% in the online gambling subforum, while total posts decreased by 63.27% in the terrestrial gambling subforum, and by 98.78% in the problem gambling subforum. In contrast, the number of posts decreased during the second lockdown period in the online (by 18.81%), in the terrestrial (by 88.66%) and in the problem gambling subforum (by 36.65%) compared to the corresponding second pre-lockdown periods (see Table 1 and Figure 3C-E).

**Table 1:** Mean number of posts and mean post length for each phase, for the entire forum and for the online, terrestrial and problem gambling subforums, respectively.

|  |  | Entire forum | OG subforum | TG subforum | PG subforum |
| --- | --- | --- | --- | --- | --- |
| $N$ posts | Pre-lockdown 1 | 7628 | 4836 | 98 | 246 |
|  | Lockdown 1 | 9489 | 6842 | 36 | 3 |
|  | Pre-lockdown 2 | 10783 | 7981 | 194 | 161 |
|  | Lockdown 2 | 8737 | 6480 | 22 | 102 |
| $M$ post length<br>(words) | Pre-lockdown 1 | 49.91 | 39.27 | 118.55 | 147.78 |
|  | Lockdown 1 | 40.25 | 35.09 | 69.44 | 15.33 |
|  | Pre-lockdown 2 | 46.96 | 43.71 | 72.38 | 56.58 |
|  | Lockdown 2 | 43.76 | 40.75 | 39.73 | 69.55 |

Note. OG: online gambling, TG: terrestrial gambling, PG: problem gambling.

**Figure 3:**
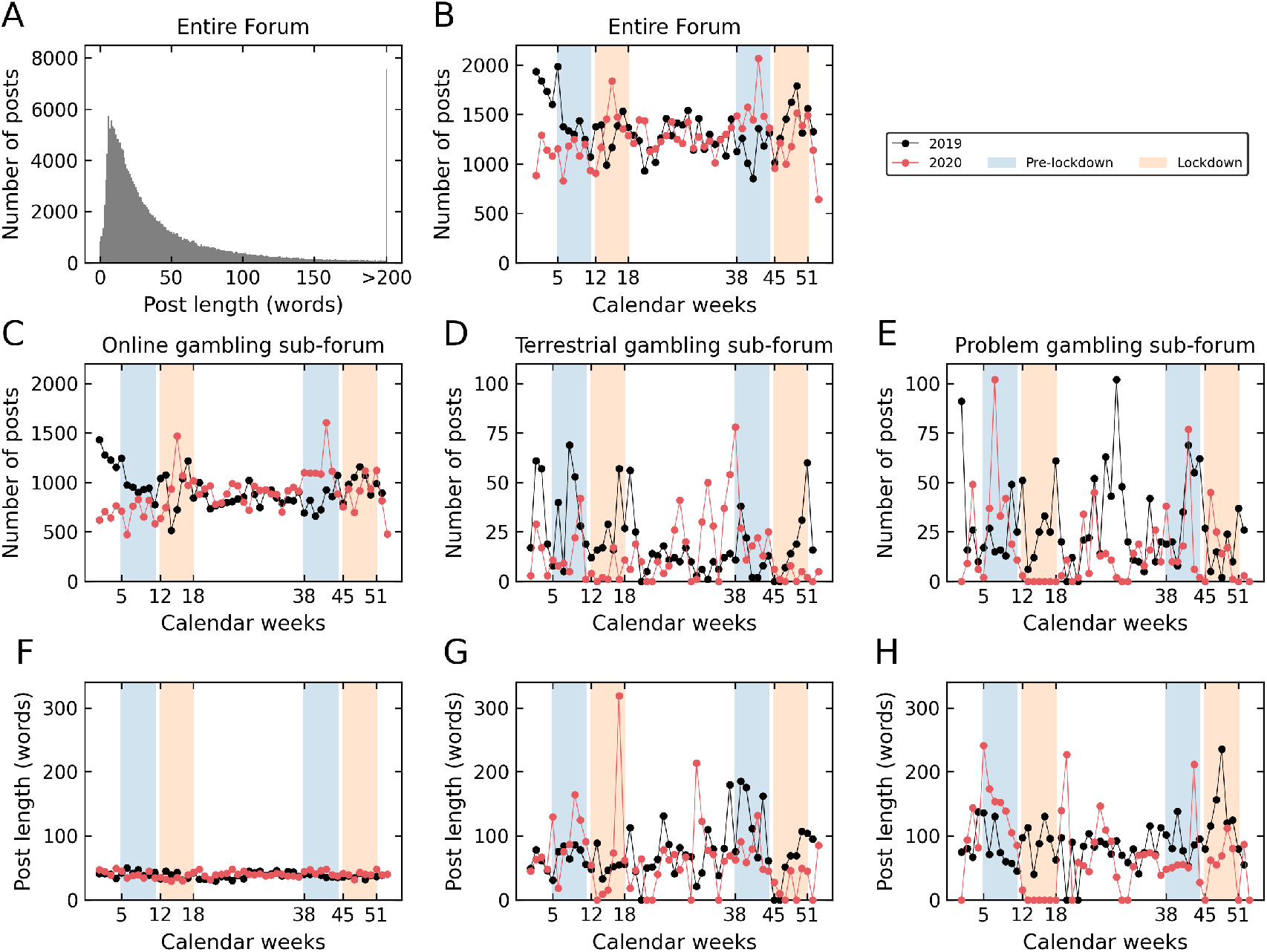
Distribution of posts for the entire forum and for the subforums. A: Distribution of post length and number of posts; B: Number of posts per year, calendar week and phase (pre-lockdown and lockdown) for the entire forum; C-E: Number of posts per year, calendar week and phase (pre-lockdown and lockdown) for the online gambling, terrestrial gambling and problem gambling subforum, respectively; F-H: Post length per year, calendar week and phase (pre-lockdown and lockdown) for the online gambling, terrestrial gambling and problem gambling subforum, respectively.

With regard to post length, on average (across all subforums), a post contained 50.12 words (*SD* = 76.44). Most posts contained 6 words and half of the posts were at least 27 words long (see Figure 3A). Posts in the terrestrial gambling subforum were longer (*M* = 81.89, *SD* = 112.93) than in the online gambling subforum (*M* = 43.75, *SD* = 65.28). The problem gambling subforum contained, on average, the longest posts among all superordinate board topics (*M* = 100.73, *SD* = 127.78). During both the first lockdown compared to pre-lockdown period, post length decreased in all three types of subforums (see Table 1). However, while the post length decreased only slightly in the online gambling subforum (reduction of 10.64%), the average post length dropped markedly in the terrestrial gambling subforum (by 41.43%) and problem gambling subforum (by 89.63%). For the second lockdown compared to pre-lockdown phase, average post length dropped by 6.77% in the online forum, and by 45.11% in the terrestrial forum, but increased by 22.92% in the problem gambling subforum (see Table 1 and Figure 3F-H).

### 3.2 Reply latencies

For each of the phases, we modelled the relationship between reply frequency and latency, respectively, between initial posts and the replies that followed as an exponential decay function with offset *y0*, asymptote *a* and decay rate *k* (see Section 2.5). To this end, the reply latencies (elapsed time since initial post) were binned into bins of 8 hours. The data and the modelled latency curves per phase (pre-lockdown and lockdown phase 1 in spring 2020, and pre-lockdown and lockdown phase 2 in winter 2020) are depicted in Figure 4. The posterior distributions of the exponential decay model parameters are depicted in Figure 5. The medians of the group-level posterior distributions were as follows: pre-lockdown 1: *y0* = 1192.55, *a* = 49.79, *k* = 1.16; lockdown 1: *y0* = 1345.42, *a* = 73.17, *k* = 1.28; pre-lockdown 2: *y0* = 1639.89, *a* = 104.05, *k* = 1.25; lockdown 2: *y0* = 1410.60, *a* = 60.75, *k* = 1.42. The medians and highest posterior density intervals of the difference distributions for the first and second phase for each of the three decay model parameters are listed in Table 2.

**Figure 4:**
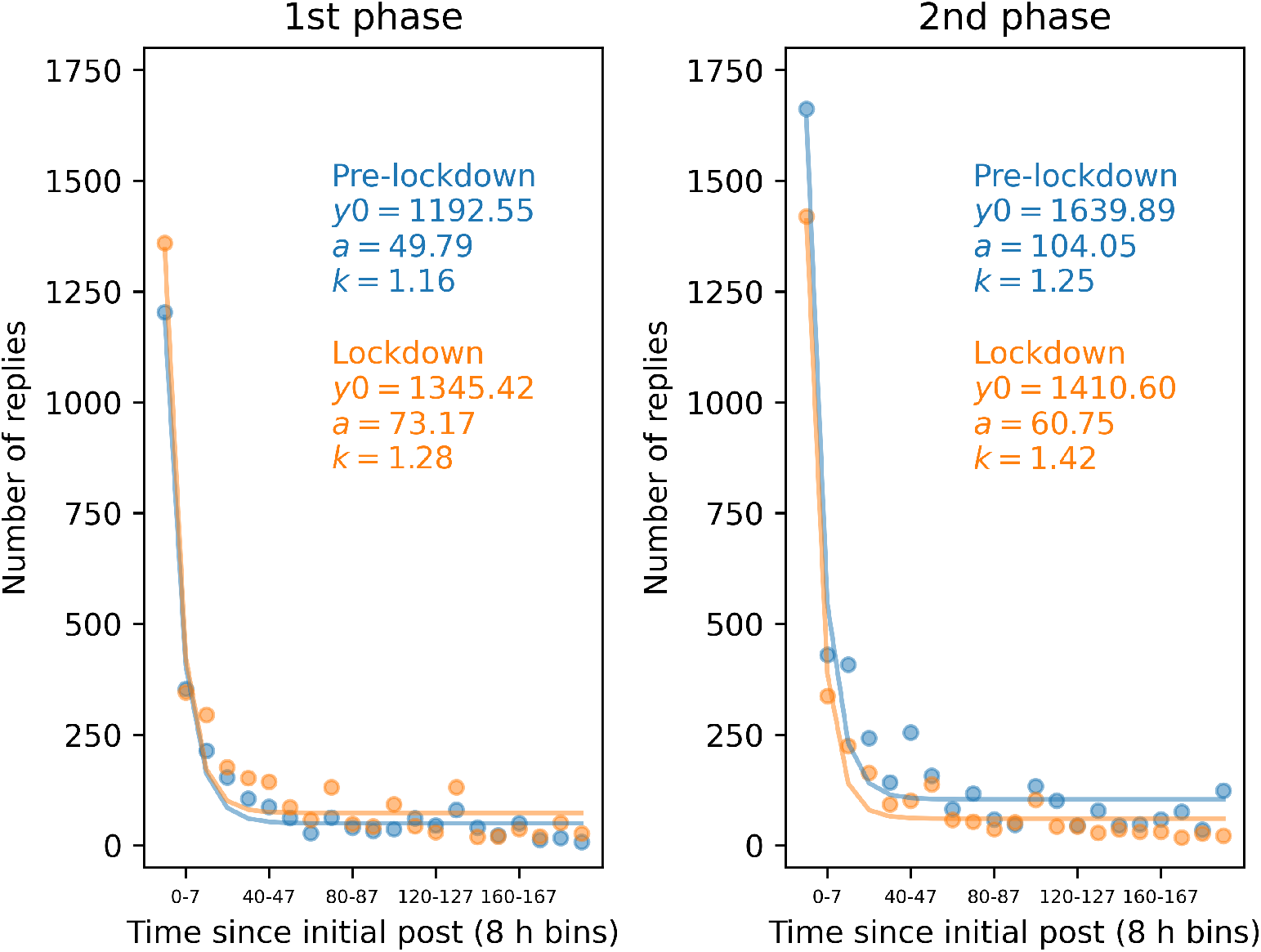
Modelled reply latency curves for the 4 phases overlayed onto the data. Reply latency (time since initial post) was binned into bins of 8 h. The latencies were modelled as exponential decay with *y0* : intercept, *a*: asymptote, and *k* : decay rate. Blue: pre-lockdown phases, orange: lockdown phases.

**Figure 5:**
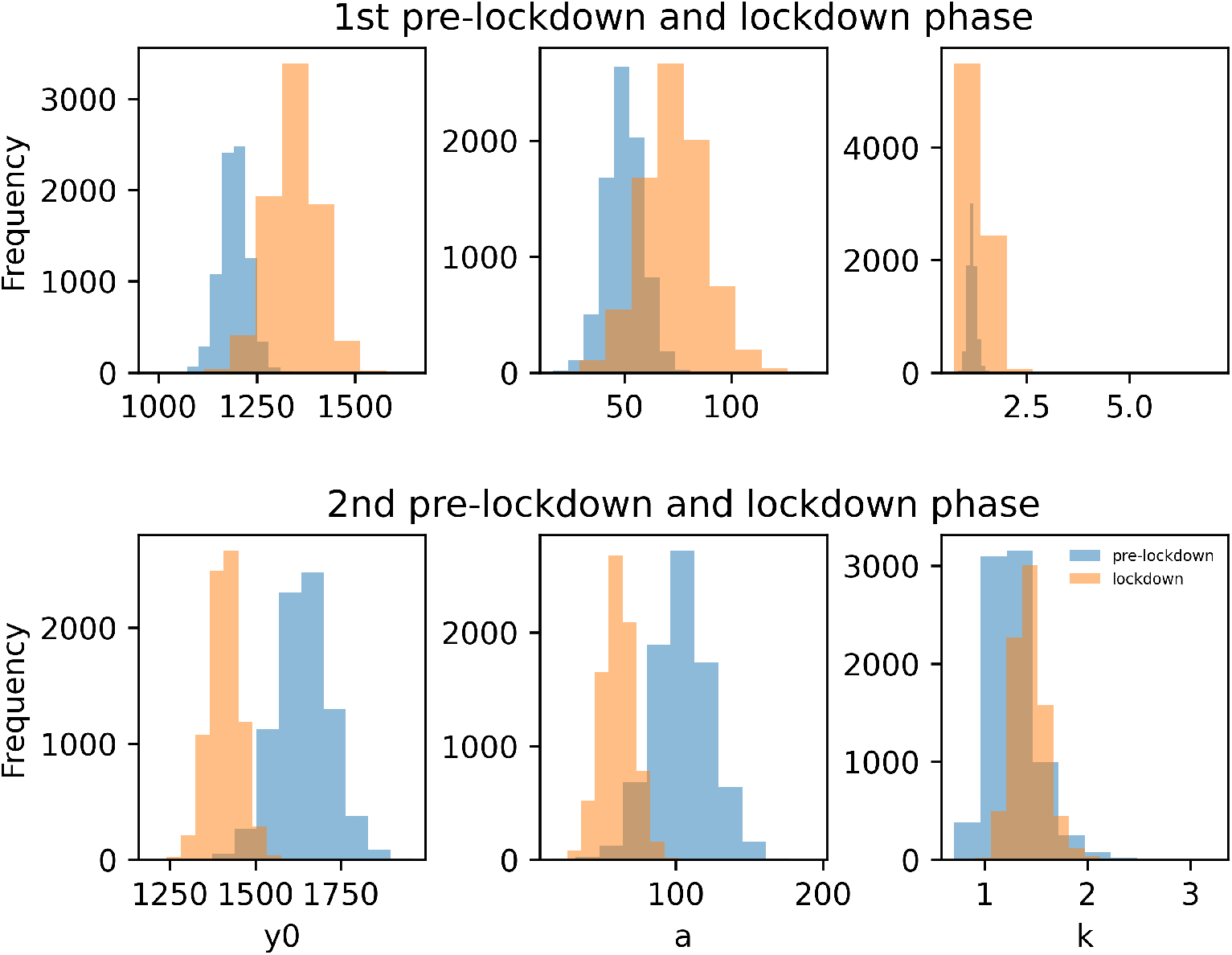
Posterior distributions of the latency model parameters for the pre-lockdown (blue) vs. lockdown (orange) phases. The latencies were modelled as exponential decay with *y0* : intercept, *a*: asymptote, and *k* : decay rate.

**Table 2:** Medians and 95% HDIs of the posterior difference distributions (lockdown minus pre-lockdown phase) for the parameters of the latency model.

|  | Phase 1 (lockdown 1 - pre-lockdown 1) | Phase 2 (lockdown 2 - pre-lockdown 2) |
| --- | --- | --- |
| $y0$ | 153.08 (17.89 to 298.24) | -230.86 (-408.76 to -44.28) |
| $a$ | 23.41 (-8.17 to 59.46) | -43.17 (-88.85 to -0.89) |
| $k$ | 0.12 (-0.30 to 0.61) | 0.17 (-0.44 to 0.69) |

*Note.* HDI: highest density interval, $y0$ : intercept, $a$ : asymptote, $k$ : decay rate.

While the intercept *y0* and the asymptote *a* were higher during the 1st lockdown compared to the 1st pre-lockdown phase, the opposite was true for the 2nd phase. Here, *y0* and *a* were lower for the lockdown compared to pre-lockdown period. The decay rate *k* was higher for both lockdown compared to pre-lockdown phases. An increase during the first lockdown compared to pre-lockdown phase was 63 times more likely for the offset *y0* (*dBF* = 62.67), 10 times more likely for the asymptote *a* (*dBF* = 10.47), and 3 times more likely for the decay parameter *k* (*dBF* = 3.37). Comparing the second lockdown compared pre-lockdown phase, a decrease was 132 times more likely than an increase for the offset *y0* (*dBF* = 132.74), and 38 times more likely for the asymptote a (*dBF* = 38.91), while an increase was 3 times more likely than a decrease for the decay rate *k* (*dBF* = 3.23).

This indicates that the number of short-latency replies was higher for lockdown phase 1 compared to pre-lockdown phase 1, while the reverse was true for lockdown phase 2 compared to pre-lockdown phase 2. In the days following an initial post, the number of replies remained higher during lockdown phase 1 compared to pre-lockdown phase 1. Here, too, the reverse pattern emerged for lockdown phase 2 compared to pre-lockdown phase 2. The decline in the number of replies was similar for phases 1 and 2.

## 4 Discussion

Using web scraping, we analysed the posting behaviour in a large German online gambling forum and changes related to the casino closures in spring and winter of 2020 that were part of the nationwide restrictions to prevent the spread of COVID-19. We hypothesised that the restrictions and their effects would be reflected in the quantitative properties of the posts, such as posting frequency and distribution across the subforums.

### Description of the discussion board

Being part of an online casino and gambling website, the main topic of the discussion board is online gambling. The online gambling subforum makes up two-thirds of all forum posts, while the problem and terrestrial gambling subforum contain only a minority of all posts. However, the online subforum has the shortest posts, followed by the terrestrial gambling subforum. The problem gambling subforum contains the longest posts, which are on average twice as long as the average post. There is a substantial number of users who write a single post (27.42% of all users), or several posts within a day, but then do not continue posting (16.96% of all users). However, many users of the forum actively post over a longer period of time (*>* 35% of all users). The classification of user types shows that almost half of the users are active in the online gambling subforum only. Only a very small percentage of users posts exclusively in the problem gambling subforum or terrestrial gambling subforum. It is particularly striking that there was a marked increase in newly registered users during the first lockdown phase in 2020 compared to the weeks preceding the lockdown. Even if it is not possible to directly infer actual gambling behaviour from participation on online forums, however, this may well reflect a lockdown-induced shift from terrestrial to online gambling.

The analysis of cross-posting behaviour of users across the three subforums examined (online, terrestrial, and problem gambling) revealed further interesting insights. Previous survey studies revealed that participation in multiple forms of gambling is linked to a greater number of problem gambling symptoms (Phillips, Ogeil, Chow, & Blaszczynski, 2013). Our cross-posting analysis is in line with these earlier observations. Only a very small minority of users posting exclusively in the online gambling subforum posted at least once in the problem gambling subforum (*N online only + PG* = 32, 1.65% of all online only users). In contrast, a substantially greater number of users classified as *mixed*, that is, users posting in both the online and terrestrial forum, contributed at least once to the problem gambling subforum (*N mixed + PG* = 195, 62.10% of all *mixed* users). While this could reflect a greater incidence of problem gambling symptoms in the at-risk *mixed* group, alternative explanations are likewise conceivable. For example, *mixed* users might simply constitute a more active subgroup, which generally participate in more subforums.

### Lockdown effects

During the first lockdown phase in spring 2020, compared to the previous reference period, there was a substantial increase in newly registered users, which likely mirrors the general increase in social media and internet usage during lockdown reported in different samples from various countries (Gupta, Jagzape, & Kumar, 2021; Lemenager et al., 2021; Vall-Roqué, Andrés, & Saldaña, 2021). Further, there was an increase in the number of posts in the online gambling subforum with a concurrent decrease in the number of posts in the terrestrial gambling subforum. This shift from terrestrial to online likely reflects the effects of the casino closures that were part of the nationwide restrictions in Germany. Data from UK samples also show that the decrease in terrestrial gambling coincided with proportionate increases in online gambling (Close et al., 2022; Emond, Nairn, Collard, & Hollén, 2022). Among the Swiss casino players who gambled during lockdown, gambling intensity decreased but online gambling increased significantly (Lischer et al., 2021).

Modelling the relationship between reply frequency and latency between initial posts and the replies that followed for each of the phases allowed us to quantify the immediacy of user engagement with posts. This revealed that the number of short-latency replies (i.e. replies that were posted within seven hours after the initial post) was substantially higher during lockdown 1 compared to pre-lockdown 1. Furthermore, throughout the week following an initial post, the number of replies remained higher during lockdown 1 compared to pre-lockdown 1, as reflected in an elevated asymptote of the decay model. However, the inverse pattern emerged during the second lockdown. Here, the number of short-latency replies was lower and also remained lower throughout the following week, as compared to the pre-lockdown period. The increase in reply latencies during the first lockdown may reflect the general marked increase in screen time and/or usage of online platforms and media after the onset of the COVID-19 pandemic (Lemenager et al., 2021; Meier et al., 2021; Pandya & Lodha, 2021).

Interestingly, this pattern was reversed during the second lockdown. The changes in the number and length of posts are also less conclusive for the second lockdown versus the corresponding reference period than for the first lockdown versus its reference period. In contrast to the first lockdown, the number of posts in the online gambling subforum decreased during the second lockdown compared to the reference period. However, “baseline” numbers in the pre-lockdown phase were higher for the second compared to the first lockdown phase. Before the first lockdown in 2020, terrestrial gambling venues were likely visited as usual. However, the degree to which this was the case prior to the second lockdown period is unclear. While gambling venues were open during this time, social distancing measures were nonetheless in place, and players may have adapted their gambling behaviour due to the infection risk associated with terrestrial gambling. However, the number of posts in the terrestrial gambling subforum increased after the first lockdown and decreased again during the second pre-lockdown phase. Nonetheless, we cannot conclude from the data whether this actually reflects increased revisiting of gambling venues after the first lockdown. It would also be conceivable to have few visits being discussed very extensively in the forum. Also, the second lockdown had been announced for some time and was foreseeable, whereas the first lockdown was a condition never experienced before. Therefore, the second pre-lockdown phase is likely affected by a variety of additional factors which complicate a direct comparison of the first and second lockdown effects. Specifically, the first pre-lockdown phase is likely a baseline that reflects “normal” gambling behaviour much more accurately than the second pre-lockdown phase. It is also not clear whether and when this “normal” situation will occur again or whether the situation before the onset of COVID-19 will be representative for the future.

Our analysis also examined posting behaviour in the problem gambling subforum, which might provide insights into potential lockdown-related effects on disordered gambling. It is particularly striking that the absolute number and length of posts in the problem gambling subforum dropped substantially during the first lockdown compared to the reference period. Perhaps problem gambling-related topics, such as relapse, recent problem gambling behaviour, tips on how to quit gambling as well as experiences with therapy, were less important for users to discuss about in the forum during the first lockdown. This change may be explained by data from an online survey from a German sample which found that most respondents gambled less or stopped gambling during lockdown, while only a minority reported increased gambling behaviour (Georgiadou et al., 2022). The results of several international studies suggest that only a small proportion of individuals gambled more frequently during lockdown, however, these appear to be particularly vulnerable individuals (Håkansson, 2020; Hodgins & Stevens, 2021; Price, 2020). However, while the number of posts in the problem gambling subforum decreased during the first lockdown, the number of posts in the online gambling subforum increased. It is not clear whether the shift from terrestrial to online gambling reflects a reduction in problem gambling, or whether people who gamble predominantly online seek less help. Online gambling was found to be more likely among persons who gamble with existing problems than among persons without gambling-related problems (Gainsbury, Russell, Wood, Hing, & Blaszczynski, 2015). Results from a study assessing an Australian sample suggest that among problem gamblers, help-seeking in online support groups or discussion boards was lower among online compared to terrestrial gamblers (Hing, Russell, Gainsbury, Blaszczynski, et al., 2015).

Addiction and COVID-19 have been described as colliding pandemics (Dubey et al., 2020; Volkow, 2020). The social distancing and lockdown measures create special challenges to individuals at risk. In the case of gambling, the closure of arcades and casinos may have led to a shift from terrestrial towards online gambling. In contrast to the terrestrial arcade, where an arcade supervisor carries out age checks and is required to identify problem gambling behaviour, online gambling is not subject to such checks. This may pose a special threat to individuals at risk for gambling addiction. The current analyses help to identify potential effects of the lockdown periods on gambling behaviour. It is highly likely that the lockdown periods have had a negative impact on the mental health of individuals at risk. The potentially detrimental effects of the pandemic and the lockdown measures on mental health, including addiction and gambling behaviour, should be monitored and may require special public health measures.

### Limitations, strengths and ethical considerations

A major limitation of our approach is that the degree to which changes in the posting behaviour reflect changes in actual gambling behaviour is unclear. A higher posting frequency, a higher mean post length and shorter reply latencies in the online gambling subforum indicate that users write more and longer posts about online gambling and respond faster to these topics. This may reflect increased online gambling behaviour. Similarly, changes in posting frequency, post length and the reply latencies in the problem gambling subforum do not necessarily represent changes in actual problem gambling behaviour or changes the prevalence of disordered gambling among forum users. Although the participation in online gambling communities is linked to problem and pathological gambling (Sirola et al., 2018, 2019), it remains unclear whether changes in participation, i.e. posting behaviour, are linked to changes in problem gambling. Linking actual gambling behaviour and selfreport represents an important avenue for future research. There is evidence that the self-reported gambling behavior is not reliable, for instance due to misestimation of losses (Heirene, Wang, & Gainsbury, 2021). Finally, the users of the forum are likely not representative for the population of all gamblers. In a Finnish sample, more than half of the people who visit online gambling communities at least once were individuals at-risk or likely pathological gamblers (Sirola et al., 2018), while the proportion is markedly lower in the general population of gamblers (Floros, 2018).

A strength of our approach lies in its high ecological validity. We examined actual posting behaviour in a real-life setting, namely posting behaviour in an online gambling forum. Self-reports are subject to several biases and limitations, such as the social-desirability bias (Krumpal, 2013) and recall bias (Bradburn et al., 1987). Posts are not self-reports in a strict sense, however, how people communicate in such forums depends, among other things, on the degree of perceived anonymity and the technical identifiability of the users. Depending on this, different norms are followed with regard to content and conversational style. This is formulated in the Social Identity Model Of Deindividuation (SIDE) model (Reicher, Spears, & Postmes, 1995). Further, there appears to be a general tendency to present oneself idealised online, in terms of psychological characteristics, which may also be reflected in the style of communication in the forum (Zimmermann, Wehler, & Kaspar, 2022). These issues may be an outlook for more specific analyses.

An advantage of using data scraped from public web forums is that large datasets may be collected without researchers’ interference, that is, there are no effects of observation. However, this also raises important ethical issues. Eysenbach and Till (2001) emphasise the importance of “perceived privacy” in an online community. If, for instance, posts can only be viewed following registration, this suggests that users might perceive their contributions as occurring in a private space (Eysenbach & Till, 2001; Landers et al., 2016). Likewise, if providers put measures in place to restrict data access, researchers should not circumvent these measures (Landers et al., 2016). We have followed these guidelines and collected data that were publicly available and did not bypass any technical barriers in doing so.

### Perspectives

In addition to a quantitative analysis of posting behaviour, a qualitative analysis of post content may yield further insight into gambling behaviour and gambling-related issues among gamblers. Comparing the subforums with regard to post length, the problem gambling subforum contained the longest posts. In many of the initial posts, users described gambling-related problems and mental health issues in great detail (“Lost everything due to addiction, fiancée, all possessions, savings turned into 25,000 euros in debt. I lose jobs all the time […]”, “Now I am faced with nothing, I am mentally breaking down, have suicidal thoughts from time to time but I would never do that !!!”, English translation). Reply posts included tips on coping with gambling addiction and discussions between users about coping strategies and treatments (“Can only advise you to stop immediately, in the end you are always the loser.”, “On the other hand, you could also use the therapy as a chance, but one that will only work if you commit yourself to it.”, English translation). Further, posts in the terrestrial gambling subforum were longer than posts in the online gambling subforum. Here too, a qualitative analysis may yield interesting insights. It is conceivable that when describing land-based gambling activities, more words are used to describe the experience and specific contextual factors (such as location, structure of the location, type of machine, etc.) in greater detail to other users. In addition, offline there may be more interpersonal communication (e.g. with other players on site) as subject of the posts.

Complementing analyses of the posting behaviour, automated text analysis and computational methods may be useful tools for identifying user topics and signs of problematic gambling behaviour (Haefeli, Lischer, & Haeusler, 2015; Hwang, Kim, Choi, Lee, et al., 2020; Maupomé, Armstrong, Rancourt, Soulas, & Meurs, 2021; McGarry & McDonald, 2017). This may facilitate player protection through early detection of gambling addiction risk, complementing human assessment, and on the part of gambling operators, prioritising customer contacts based on risk assessment (Haefeli et al., 2015).

## Data Availability

All data with the exception of data related to individual user profiles and user names are openly available at https://osf.io/rwym3/.

## Author contributions

**Conceptualisation:** J. P.

**Methodology:** E. S., K. H. K. K., J. P.

**Software:** E. S., S.M.

**Resources:** E. S., J. P.

**Data Curation:** E. S.

**Formal analysis:** E. S., S. M.

**Writing - Original Draft:** E. S., S. M.

**Writing - Review & Editing:** E. S., S. M., K. H. K. K., K. K., N. R., J. P.

**Visualisation:** E. S., S. M.

**Project administration:** J. P.

**Supervision:** J. P., N. R.

## Supplement

**Table S 1:** Casino closure periods in the federated states of Germany.

| State | 1st lockdown period |  | 2nd lockdown period |  |
| --- | --- | --- | --- | --- |
|  | Start | End | Start | End |
| Baden-Württemberg | 17/03/2020 | 10/05/2020 | 02/11/2020 | 07/06/2021 |
| Bavaria | 17/03/2020 | 10/05/2020 | 02/11/2020 | 07/06/2021 |
| Berlin | 14/03/2020 | 01/06/2020 | 02/11/2020 | 18/06/2021 |
| Brandenburg | 18/03/2020 | 27/05/2020 | 02/11/2020 | 11/06/2021 |
| Bremen | 18/03/2020 | 26/05/2020 | 02/11/2020 | 31/05/2021 |
| Hamburg | 15/03/2020 | 26/05/2020 | 02/11/2020 | 04/06/2021 |
| Hessen | 18/03/2020 | 14/05/2020 | 02/11/2020 | 10/06/2021 |
| Mecklenburg-Western Pomerania | 18/03/2020 | 14/06/2020 | 02/11/2020 | 07/06/2021 |
| Lower Saxony | 17/03/2020 | 24/05/2020 | 02/11/2020 | 31/05/2021 |
| North Rhine-Westphalia | 16/03/2020 | 10/05/2020 | 02/11/2020 | 28/05/2021 |
| Rhineland-Palatinate | 18/03/2020 | 26/05/2020 | 02/11/2020 | 02/06/2021 |
| Saarland | 18/03/2020 | 03/05/2020 | 02/11/2020 | 31/05/2021 |
| Saxony | 19/03/2020 | 14/05/2020 | 02/11/2020 | 14/06/2021 |
| Saxony-Anhalt | 18/03/2020 | 27/05/2020 | 02/11/2020 | 25/05/2021 |
| Schleswig-Holstein | 14/03/2020 | 17/05/2020 | 02/11/2020 | 31/05/2021 |
| Thuringia | 18/03/2020 | 12/05/2020 | 02/11/2020 | 02/06/2021 |
| Overlap | 19/03/2020 | 03/05/2020 | 02/11/2020 | 25/05/2021 |

